# Higher Relative Viral Load Excretion Determined by Normalised Threshold Crossing Value in Acute Cases infected with the B.1.1.7 Lineage VOC 202012/01 (Using S gene target failure as a Proxy) When Compared to other Circulating Lineages in Wales

**DOI:** 10.1101/2021.04.02.21254832

**Authors:** Anastasia Couzens, Isa Murrell, Laura Gifford, Ben Johns, Kathleen Pheasant, Luke Turner, Jonathan Evans, Catherine Moore

**Affiliations:** Wales Specialist Virology centre, Public Health Wales Microbiology Cardiff, University Hospital of Wales, Heath Park, Cardiff, CF14 4XW

## Abstract

Since the emergence of SARS-CoV-2, global monitoring of the virus using whole genome sequencing has identified mutations occurring across the viral genome. Whilst the majority have little impact on the virus, they are used effectively to monitor the movement of the virus globally and to inform locally on transmission chains.

In late 2020, a variant of SARS-CoV-2 (B.1.1.7 - VOC 202012/01) was identified in the UK with a distinct constellation of mutations, including in the spike gene that increased transmissibility. A deletion in spike also affected one of the screening qPCR tests being used in the UK outside of Wales, causing a failure to detect the target. This quickly became a surrogate marker for the variant to allow rapid monitoring of the virus as it seeded into new regions of the UK.

A screening study using this assay as a proxy marker, was undertaken to understand the prevalence of the variant in Wales. Secondary analysis of a screening qPCR that didn’t target the S gene and also included an endogenous control, was also performed to understand viral load excretion in those infected with the variant compared to other circulating lineages. Using a combination of analytical methods based on the C_t_ values of two gene targets normalised against the endogenous control, there was a difference in the excreted viral load. Those with the variant excreting more virus than those not infected with the variant. Supporting not only increased infectivity but offering a plausible reason why increased transmission was associated with this particular variant.

Whilst there are limitations in this study, the method using C_t_ as a proxy for viral load can be used at the population level to determine differences in viral excretion kinetics associated with different variants.

## Introduction

Severe acute respiratory syndrome coronavirus (SARS-CoV-2) was first detected within the United Kingdom (UK) in January 2020, with the first confirmed case identified in Wales at the end of February 2020 (Welsh Government, 2020; Lillie *et al*., 2020). Variants of viruses commonly arise due to mutation in the viral genome, and are mostly identified via whole-genome sequencing (Burki, 2021). Mutations and variants are often of little consequence with little effect on the virus or naturally burn-out over time, however can become concerning if the change causes a competitive advantage over the ancestral phenotype (Yurkovetskiy *et al*., 2020). A variant of lineage B.1.1.7 (1) was first identified in the South East of England, and was designated Variant of Concern (VOC) 202012/01 in December 2020 (PHE, 2020). Preliminary analysis suggested an approximate growth rate 71% higher than other variants, and reported estimated increase of between 40% and 70% in transmissibility (ECDC, 2020; NERVTAG, 2020).

VOC 202012/01 is characterised by an unprecedented number of mutations including an out of frame six nucleotide deletion causing the removal of histidine and valine at positions 69 and 70 respectively (ΔH69/V70) from the S1 protein subunit (Rambaut *et al*., 2020; Gloubchik *et al*., 2021). In total there are eight lineage-defining amino acid changes in Spike, in addition to pre-existing D614G, which are of particular concern due to the antigenicity of the peplomer (Rambaut *et al*., 2020; Yurkovetskiy *et al*., 2020).

‘Lighthouse laboratories’ provide mass testing for ‘Pillar 2’ of the UK government’s ‘5-pillar plan’ for the upscaling of SARS-CoV-2 testing, whereas the Public Health England and Wales (PHE/PHW) labs and National Health Service (NHS) hospital laboratories cover ‘Pillar 1’ (UK Department of Health and Social care, 2020). In 3 of these Lighthouse laboratories, a phenomenon of S-gene target failure (SGTF) was noticed in Applied Biosystems™ TaqPath™ SARS-CoV-2 qualitative reverse transcriptase polymerase chain reaction (RT-qPCR) assay (PHE, 2020). SGTF was found to indicate the presence of VOC 202012/01 in samples due to ΔH69/V70, with further study showing that, in samples where both sequence and SGTF status are known, 99.3% of samples with ΔH69/V70 sequences are SGTF; furthermore, 98% of all ΔH69/V70 samples in England were identified as being VOC202012/01 by the beginning of week 50/2020 (PHE, 2020). With this reasoning, the TaqPath™ assay was used as a supplementary test in PHW’s laboratory in University Hospital of Wales (UHW) on samples that had tested positive via other RT-PCR assays from across Wales in order to provide a crude screen for VOC 202012/01 in Pillar 1 samples. An incidental observation of such was the ability to compare original cycle threshold (C_t_) values of samples with and without SGTF using both Livak and Schmittgen’s ΔΔC_t_ comparative method (2001), and a relative quantification method in which the ratio of target gene C_t_ compared to internal control (IC) C_t_ was calculated. Using SGTF as a crude indicator of the presence of VOC 202012/01 and representing a unique insight into Pillar 1 testing, this analysis sought to determine if there is a statistically significant difference in the relative quantification of virus, and in the relative viral gene expression in VOC 202012/01 vs non-VOC 202012/01 when normalised C_t_ values obtained by independent means are used as a proxy in lieu of viral loads.

## Methods

Between the 21^st^ December 2020 and 26^th^ January 2021, all dry oropharyngeal/nasopharyngeal swabs from across Wales that returned a positive result. These samples were split into those with C_t_ values of ≤27 for any gene target, and those with C_t_ values >27 for all gene targets, as the Pathogen Genomics Unit of UHW were to have priority access to samples with C_t_≤27 for pre-existing whole-genome sequencing efforts.

Samples were largely received as 96-deep-well extraction plates containing extracts with a mix of both positive and negative results. Positive samples were manually picked based on plate maps constructed using RT-qPCR results and stored at 5°C until further testing. Sample-specific NHS barcodes and episode numbers were utilised for record keeping of processed samples, and to allow for future clinical and epidemiological investigation using the Laboratory Information Management System (LIMS) TrakCare ^®^ Lab by InterSystems™ (Cambridge, Massachusetts, United States) used in PHW labs.

The TaqPath™ assay was performed on all sample extracts, according to kit instructions, and results were split into 3 qualitative result categories independent of the C_t_ results obtained through the original diagnostic testing method; ‘Negative’ (RNA not detected by TaqPath but originally reported as positive), ‘SGTF’ and ‘Wild-type (WT)’. These completed results were forwarded to Public Health Wales Communicable Disease Surveillance Centre for analysis to understand the wider epidemiology of SGTF in Wales.

To analyse the data further locally, those samples that were tested using the PerkinElmer ^®^ (Waltham, Massachusetts, United States) (PE) SARS-CoV-2 RT-qPCR assay, were selected due to the use of an endogenous control to normalise C_t_ values against and because S was not a target, the kinetics of the assay would likely be not affected by absence of amplification or detection in this region.

The original PCR C_t_ values for the PE Nucleocapsid (N) and Open-Reading Frame 1ab (ORF1/ab) gene targets, plus the undisclosed human-originating endogenous internal control (IC), were then recorded for each sample. Together with the corresponding TaqPath™ result, were then split into populations based on the aforementioned categories. Samples with a complete target failure of either N gene or ORF/1ab in the PE assay were then excluded from the final analysis. ΔC_t_ was then calculated for each sample by subtracting the value received for IC from the value received from the gene target.

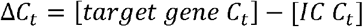

This was completed for both the N gene, and then ORF1/ab. Mean and median for these ΔC_t_ values were then calculated in order to show crude differences in averages.

Livak and Schmittgen’s (2001) ΔΔC_t_ method was conducted for each target gene; with all calculations taking place in Microsoft ^®^ (Redmond, Washington, United States) Excel ^®^ 2016 (V 16.0.5095.100). The results of which then underwent log transformations for ease of statistical analysis, and subject to one-way ANOVA and Tukey’s multiple comparisons post-hoc test in Prism ^®^ (V 9.0.1) by GraphPad Software (San Diego, California, United States). Data was also plotted for normality in order to satisfy an assumption of ANOVA. In order to perform the ΔΔC_t_ method, the C_t_ values for N gene, ORF1/ab and IC for the positive control (PC) reported for every PerkinElmer ^®^ plate tested at the UHW PHW site between 21^st^ December 2020 and 26^th^ January 2021 inclusively were recorded retroactively.

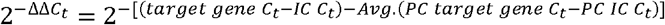

Normalisation with regards to the IC was also performed in which the ratio of target gene C_t_ to IC C_t_ was determined. Normality was plotted, and all data was subject to one-way ANOVA and Tukey’s multiple comparisons post-hoc test.

## Results

In total, 849 samples that tested positive in the PerkinElmer ^®^ assay were also tested using the TaqPath™ assay, 65 samples were excluded from the final analyses due to target failure in the PE assay. Of those analysed, 119 were ‘Negative’ according to the TaqPath™ assay, 275 were ‘SGTF’, and 390 were ‘WT’.

The mean ΔC_t_ for ‘SGTF’ samples were lower than ‘WT’ by 3.43 (3s.f.) data points for N gene, and 3.78 (3s.f.) for ORF1/ab; and lower than ‘Negative’ samples by 8.28 (3s.f.) and 10.4 (3s.f.) for N gene and ORF1/ab, respectively. Similar differences are retained for the median average, with median ΔC_t_ for ‘SGTF’ samples dwarfing those for ‘WT’ by 5.00 (3s.f.) for N gene, and 4.00 (3s.f.) for ORF1/b; and those for ‘Negative’ by 10.0 (3s.f.) and 11.0 (3s.f.) for N gene and ORF1/ab, respectively (Table 1).

**Table 1:** shows the mean and median ΔC_t_ values calculated for both N gene and ORF1/ab, as determined by PerkinElmer® SARS-COV-2 RT-qPCR. ΔC_t_ results are split into 3 groups (‘Negative’, ‘SGTF’, ‘WT’) which are based on independent qualitative results from TaqPath™ SARS-COV-2 RT-qPCR.

|  | Negative |  | SGTF |  | WT |  |
| --- | --- | --- | --- | --- | --- | --- |
| $\Delta C_t$ | N gene | ORF1/ab | N gene | ORF1/ab | N gene | ORF1/ab |
| Mean | 9.31 | 10.7 | 1.03 | 0.331 | 4.46 | 4.11 |
| Median | 10.0 | 11.0 | 1.00 | 0.00 | 5.00 | 4.00 |

The distribution of data point values, with statistical significance between groups indicated. QQ normal plots for C_t_ values compared by both the ΔΔC_t_ comparative method and relative quantification by ratio proved that both sets of data satisfied the normality (figure 1). Furthermore, all samples were independent with a continuous dependent variable so, assuming equal variance, both datasets were deemed suitable for analysis by one-way ANOVA. Results presented in Fig.1A indicated that there was a statistically significant difference in the log-fold difference relative to the IC C_t_ between each paired gene target in the ‘SGTF’, ‘WT’ and ‘Negative’ groups (F(3, 781) = 113 3s.f., *p* <0.0001).

**Figure 1:**
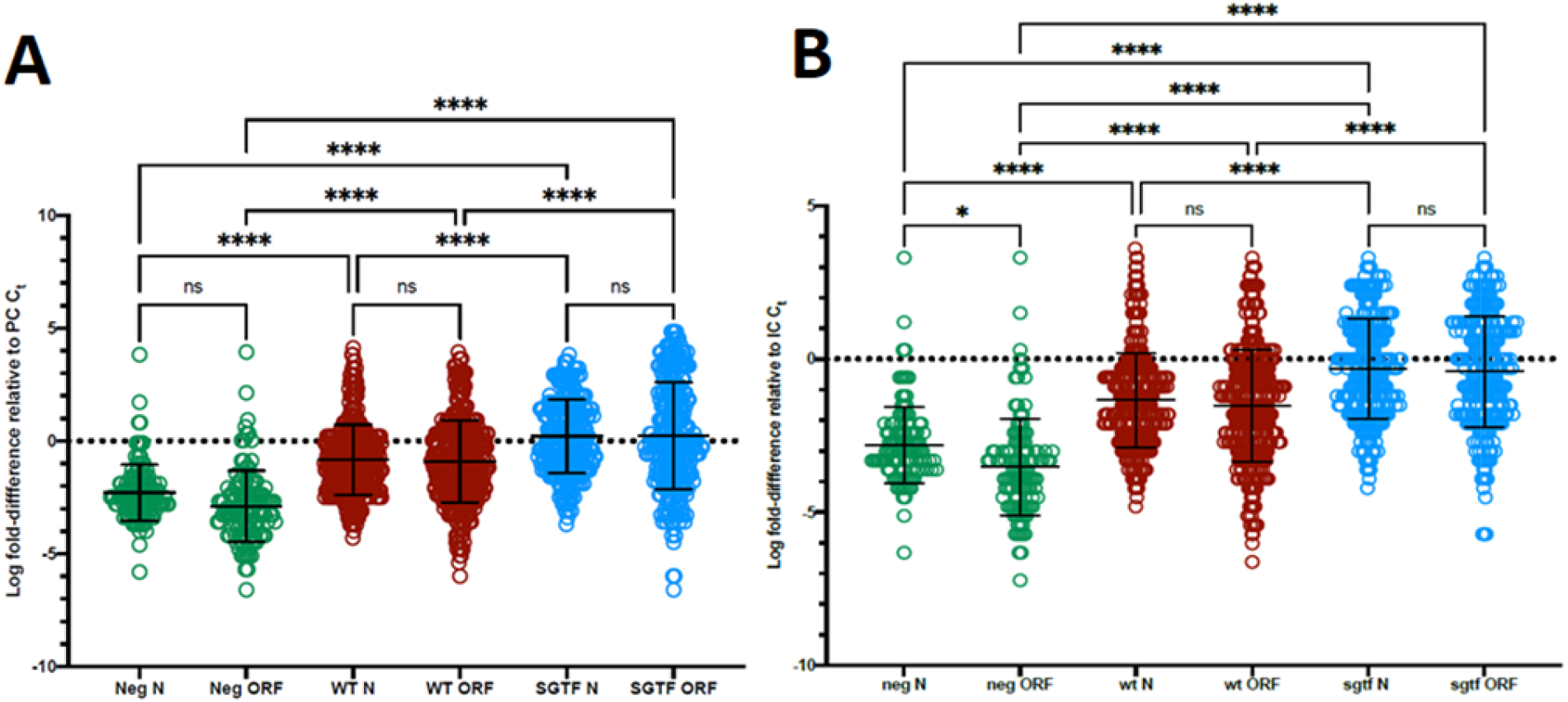
shows the distribution of data C_t_ analysed by Livak and Schmittgen’s (2001) ΔΔC_t_ comparative method for relative gene expression (Fig. 1A) and by relative quantification by ratio (Fig. 1B) for both N gene and ORF/ab for samples grouped together as ‘Negative’ (green), ‘WT’ (red) and ‘SGTF’ (blue) – determined by TaqPath™ assay. Data analysed by one-way ANOVA and Tukey post-hoc test, the degree of significance of which are indicated by asterisks, whereby 4 asterisks (****) indicate a p value <0.0001, 1 (*) indicates a p value <0.05, and ‘ns’ denotes a lack of statistical significance.

A Tukey post hoc test of multiple comparisons revealed that the relative gene expression in ‘SGTF’ samples was higher than in ‘WT’ (1.03 ± 0.401 3s.f., *p* <0.0001; 1.14 ± 0.401 3s.f., *p* <0.0001) and ‘Negative’ (−2.49 ± 0.559 3s.f., *p* <0.0001; −3.12 ± 0.559 3s.f., *p* <0.0001) samples, for N gene and ORF1/ab respectively. No statistical significance was detected between the log fold-difference relative to IC C_t_ of gene targets within the same qualitative TaqPath™ group. Similar results were observed when normalising to the IC using ratio data, shown in Fig.1B; whereby one-way ANOVA revealed a statistically significant difference in the relative quantification between each paired gene target in the 3 groups (F(3, 781) = 98.8 3s.f., *p* <0.0001). A Tukey test showed that the log-fold difference was higher in ‘SGTF’ samples than in ‘WT’ (−1.03 ± 0.374 3s.f., *p* <0.0001; −1.14 ± 0.374 3s.f., *p* <0.0001) and ‘Negative’ (−2.49 ± 0.521 3s.f., *p* <0.0001; −3.12 ± 0.521 3s.f., *p* <0.0001) samples, for N gene and ORF1/ab respectively. Furthermore statistical significance was detected in the difference between the log-fold relative C_t_ values of N gene and ORF1/ab in the ‘Negative’ group (0.718 ± 0.679 3s.f., *p* = 0.0115), and no statistically significant difference was detected between the differences in relative quantification of the paired gene targets within the remaining 2 groups.

## Discussion

These results indicate a trend in which viral target gene fold-difference relative to human endogenous IC is higher in samples that result in SGTF than in those that do not. In this analysis, rather than assuming and using the negative correlation between raw C_t_ values and viral load, normalisation of the target gene to the internal control was taken into account, which builds upon C_t_ analysis carried out in other studies (Calistri *et al*., 2021; Walker *et al*., 2021).

When using SGTF as an indicator of VOC 202012/01, and a significantly higher calculated log fold-difference relative to the IC as a proxy for viral load in these samples, the results support other studies that show patients infected with VOC 202012/01 excrete higher viral loads than patients with earlier circulating lineages of SARS-CoV-2 (Kidd *et al*., 2020; Golubchik *et al*., 2021). Supporting the suggested increased infectivity and transmissibility reported for this variant.

A limitation of this analyses is as this was performed retrospectively, the efficiency of the amplification kinetics of the PE assay was not determined. Because of this, the original iteration of the ΔΔC_t_ method in which efficiency is assumed to be 100%, was used.

Using a single assay and gene target failure to determine the presence or not of a particular variant also has limitations, as despite genomics showing that in regions where the variant might dominate 98% or more of the target failure samples are positive for the VOC, the change that leads to the failure may also be sporadic. For example, ΔH69/V70 was reported in some circulating lineages before the emergence of VOC 202012/01, In Wales however, the molecular epidemiology generated by systematic whole genome sequencing in pillar 1 and latterly in pillar 2, largely support the data generated by the proxy assay for the arrival and establishment of VOC 202012/01 across Wales.

This study does demonstrate that whilst C_t_ values generated in a qualitative PCR test can’t be used to firmly determine infectivity, they can be used to indicate changes in viral load excretion patterns associated with emerging variants at a population level, when a common testing platform with an endogenous control is used to normalise target gene against.

## Data Availability

All data will be made available by request from the corresponding author

## Acknowledgements

The laboratory staff across the Public Health Wales and NHS Wales laboratory network for providing the extracts for testing. Tom Connor, Sally Corden and the Staff of the Pathogen Genomics Unit for supporting the validation of the data generated by the SGTF screening study. Chris Williams and Clare Sawyer and the team in the PHW variant cell for feedback on the manuscript and for supporting the wider epidemiological data analysis of the SGTF screening study. Public Health Wales IMT for providing the funding to support the SGTF screening study.

